# Evolution of Reporting P-values Across the Biomedical Literature, 1990-2025: an Updated Meta-Research Study

**DOI:** 10.64898/2026.01.14.26344149

**Authors:** Jinhyeok Choi, Keeheon Lee, David Chavalarias, Jae Il Shin, John P A Ioannidis

**Author notes:** **Correspondence to:** Jae Il Shin, M.D., Ph.D., Address: 50 Yonsei-ro, Seodaemun-gu, C.P.O. Box 8044, Department of Pediatrics, Yonsei University College of Medicine, Seoul 03722, Republic of Korea, John P A Ioannidis, M.D., D.Sc., Departments of Medicine, Epidemiology and Population Health, Biomedical Data Science, and Statistics, and Meta-Research Innovation Center at Stanford (METRICS), Stanford University, Stanford, CA, USA.

## Abstract

**Importance:** Over several decades there have been extensive debates on the use and misuse of statistical significance. It would be important to capture what *P*-values are reported in biomedical papers and whether their patterns have changed over time.

**Objective:** To quantify the reporting dynamics on *P*-values in biomedical articles in PubMed and PubMed Central(PMC) database over a 35-year period (1990-2025).

**Design:** Data were retrieved from the National Library of Medicine via PubMed and PubMed Central (PMC, full-text articles included), fetching the entire accessible corpus. Records were computationally processed using a regular expression algorithm, validated for various mathematical formats, to extract reported *P*-values from the text.

**Setting:** The study includes 22,734,796 PubMed abstracts, 6,031,459 PMC abstracts and 6,397,787 PMC full-texts.

**Main Outcomes and Measures:** Proportion of article reporting at least 1 *P*-value, at least 1 *P*-value less than thresholds (.05 and .005), distribution of *P*-values by magnitude and operator type.

**Results:** The proportion of articles reporting *P*-values increased from 7.5% in 1990 to 18.3% in 2025 for PubMed abstracts, and from 5.2% to 53.3% for PMC full-texts. The median number of *P*-values per article increased from 2 to 7 in PMC full-text articles, with upward trends observed in all databases. A high proportion of *P*-values remains clustered around .05 and .001 in all databases. The proportion of articles reporting at least one *P*-value ≤ .05 has remained in the range 94%-98% since 1998, while the proportion reporting at least one *P*-value≤ .005 has increased over time, reaching 57.0% for PubMed abstracts and 62.5% for PMC full-texts. The reporting of ‘exact’ *P*-values increased until 2015, but with no further increase in the last 10 years (PubMed abstracts: 17.6% in 1990, 51.1% in 2015, 49.8% in 2025)

**Conclusions and Relevance:** Our evaluation demonstrates the pervasive entrenchment of *P*-values, despite heavy debates and major changes in the content of the biomedical literature over time. More *P*-values are reported and papers using *P*-values almost always report some that are statistically significant. Readers should remain aware of the major issues surrounding *P*-value misuse and misinterpretation.

**Key points:** *Question:* With continuing debate regarding the use and misuse of statistical significance, how have reported P-values evolved over the past 35 years?

*Findings:* Across over 22 million PubMed abstracts and over 6 million full-texts, reporting of P-values became more common over time. Almost all (94-98%) abstracts and full-text reporting P-values have at least one significant at the .05 threshold. The reporting of exact P-values increased until 2015 but plateaued since then. Clustering around traditional statistical significance thresholds remains consistent.

*Meaning:* P-values reporting has become more common over time, with pervasive prevalence of significant P-values across the biomedical literature.

## Introduction

There has been extensive debates regarding the use and misuse of P-values.^1^ In the last decade, these have included calls to redefine thresholds (e.g. use a stricter P<.005 instead of P<.05 for claiming significance) or abandon ‘statistical significance’ entirely.^2–4^ In the last decade, biomedical research has also witnessed other major transformations that may have affected whether and how P-values are reported and what reported P-values might be. These transformations include more sensitization about using effect sizes and confidence intervals rather than P-values; a large volume of high-dimensional (e.g. omics) studies with potentially lower P-values and P-value thresholds; and strengthened pleas and efforts for replication studies and reporting “negative” results.^2,5,6^ A previous meta-research study had evaluated abstracts in the entire PubMed and full-text open-access articles from PubMed Central in 1990-2015.^7^ That evaluation found that P-values were widely used and, whenever they were used, almost all abstracts and full-text articles had some statistically significant P-values based on the .05 threshold. Here, we aimed to evaluate the evolutionary trends of P-values reported in biomedical research articles extending the analyses until 2025, thus covering a period of 35 years.

## Methods

This meta-research study extends a previous evaluation using the same methods.^7^ In brief, we performed automated text-mining analysis on the entire PubMed and PubMed Central (PMC), in the period 1990-2025. While PubMed primarily provides public access to abstracts, PMC offers access to abstracts and full texts articles. For the PubMed analysis, we extracted all P-value data from MEDLINE-indexed articles with abstracts published between January 1, 1990, and September 23, 2025. The same methodology was applied to the PMC Open Access subset (downloaded October 11, 2025), analyzing both the abstracts and the full-texts. We downloaded xml files for each article that generally do not capture figures, tables, and supplements.

We constructed a P-value detection algorithm based on regular expressions, building upon the established method used in the previous evaluation with several refinements.^7^ A ‘P-value report’ was defined as a text string comprising a variation of the term ‘P-value’, followed by a relational operator and a numeric value. The complete source code is available in the Supplement.

As done previously, we separately examined specific article categories with high clinical relevance.^7^ These categories, defined by MEDLINE indexing, included randomized controlled trial, clinical trial, review, and meta-analysis. To maintain consistency with the previous study, we adopted the ‘Clinically Useful Journals’ list as a following of the NLM ‘Core Clinical Journals’ filter, which has retired in 2021.^8^ To prevent data overlap, the ‘clinical trial’ category excluded articles classified as ‘randomized controlled trial’, and the ‘review’ category excluded those classified as ‘meta-analysis’, as done previously.^7^

We evaluated the following characteristics of P-values and their trends over time: proportion of abstracts and full texts containing P-values; distribution of reported P-values; minimum (most statistically significant) and maximum (least statistically significant) P-values reported; proportion of abstracts and full texts reporting at least one P-value below the thresholds of .05 and .005; distribution of relational operators used in reported P-values; and number of P-values per paper.

Analyses were performed using Python version 3.11.12 (Python Software Foundation) and Microsoft Excel (Microsoft Corp).

## Results

From 1990 to 2025, PubMed database included 22,734,796 articles with an abstract, of which 3,500,485 abstracts reported P-values (**Figure 1A**). Proportion of abstracts reporting P-values increased annually from 7.5% in 1990 to 17.3% in 2015 and since then has remained relatively stable (18.3% in 2025). In 2025 the highest proportion was 58.3% in randomized controlled trials, followed by 41.0% in meta-analyses, 31.7% in clinically useful journals, 30.5% in clinical trials, and 1.0% in reviews. The increase over time pertains to all categories except for reviews and clinically useful journals.

**Figure 1.**
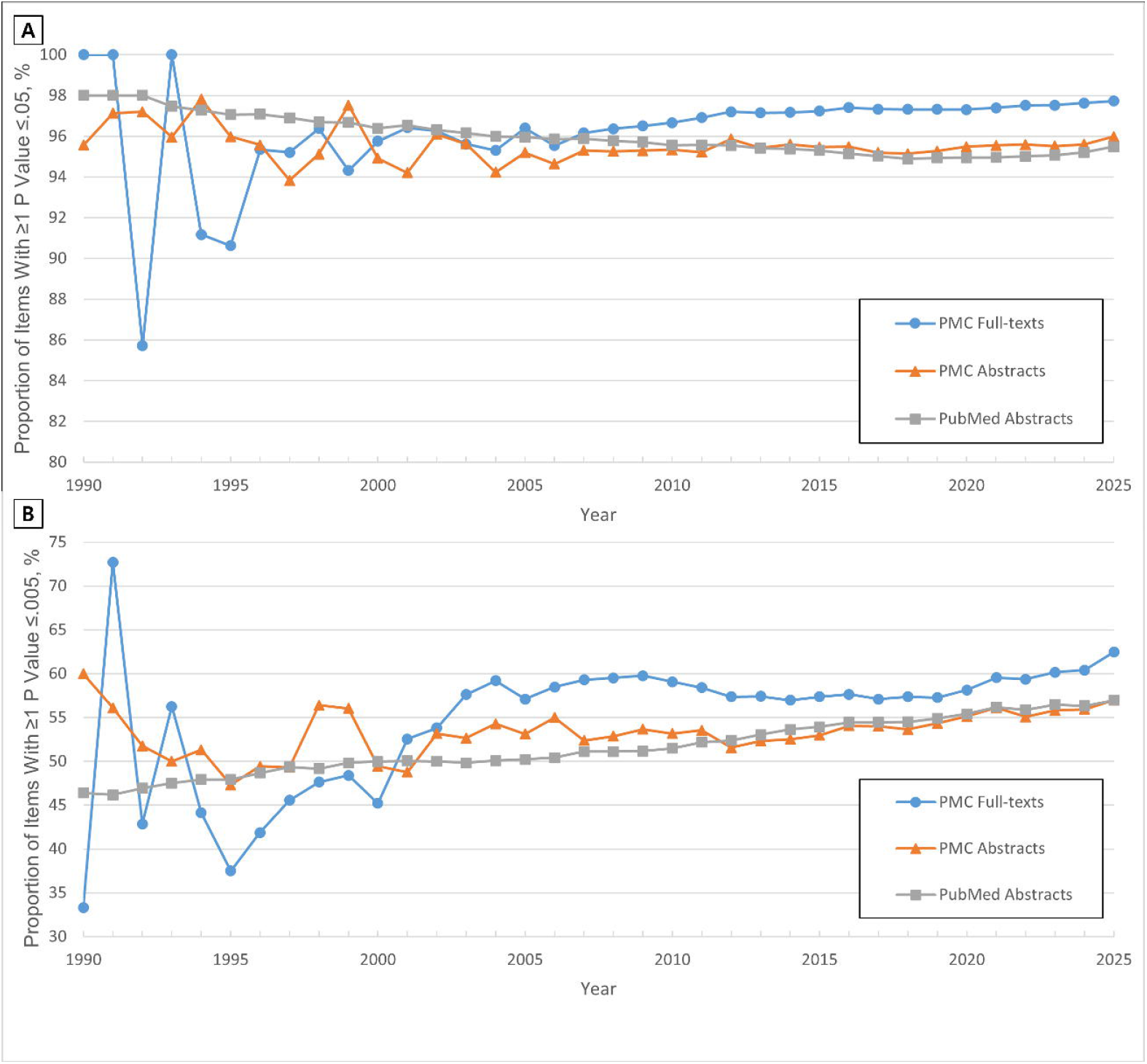
(A) Proportion of PubMed abstracts reporting at least 1 *P*-value in the period 1990-2025 for different article categories. (B) Proportion of PubMed abstracts, PMC abstracts, and PMC full-texts reporting at least 1 *P*-value in the period 1990-2025. The graph presents the annual proportion of abstracts that reported at least 1 *P-*value.

We also evaluated 6,031,459 PMC abstracts and 6,397,787 PMC full-texts. The proportion with P-values in PMC full-texts was 5% in 1990, 48.9% in 2015 and 53.3% in 2025 (**Figure 1B**).

In all databases, the distribution of P-values showed strong clustering around .05 and .001, followed by .01 (**Figure 2, Supplementary Figure 1, 2**). Operator ‘<’ was most prevalent for .001 and .05, and operator ‘=’ was most prevalent for values higher than .05.

**Figure 2.**
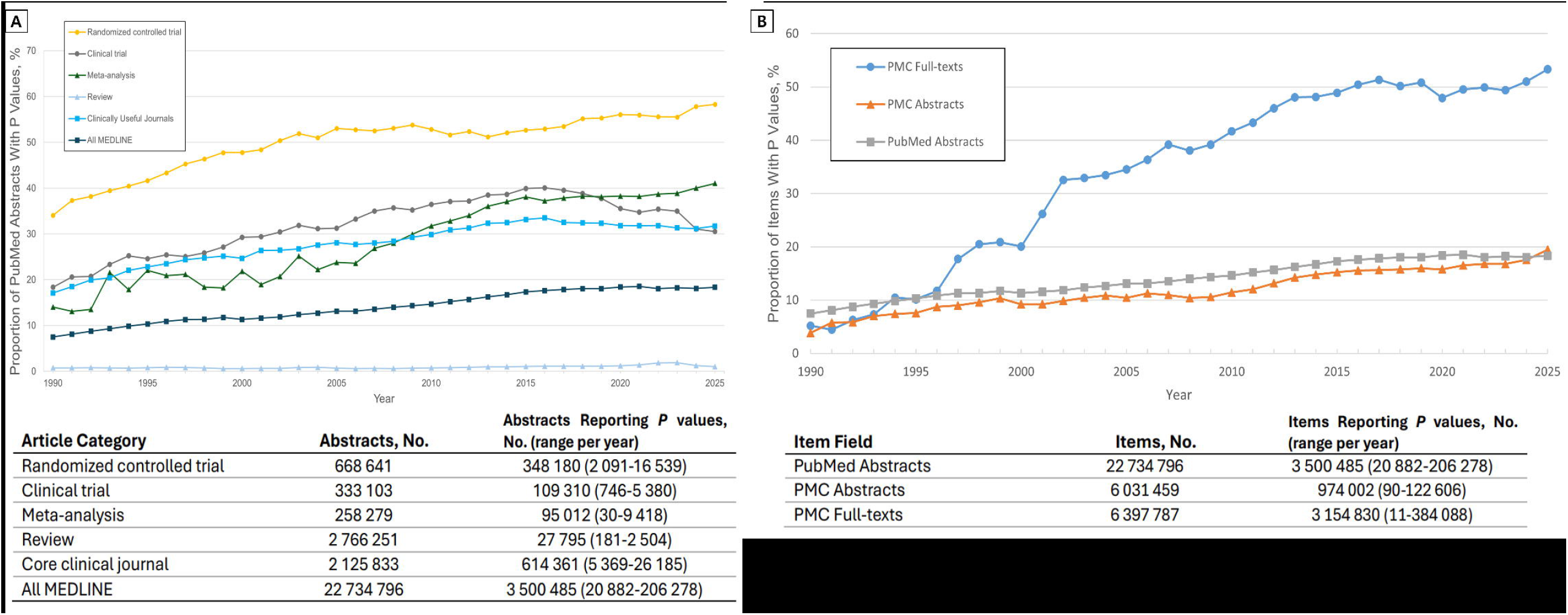
Distribution of *P*-values in 5,454,203 PMC full-texts in the period 2015-2025. There is a total of 51 bins shown, each with a width of .001, except for the rightmost bin, which represents the number of non-significant *P*-values (.05<*P*).

Mean −log_10_ of minimum P-values was largely stagnant or slightly increasing (2.33 in 1990 to 2.45 in 2025) in PubMed abstracts, was steady in PMC full-texts, and slightly decreased in PMC abstracts (2.53 in 1990 to 2.45 in 2025). Mean −log_10_ of maximum P-values decreased from 1.80 in 1990 to 1.57 in 2025 and the decrease was consistent also in PMC databases (**Supplementary Figure 3-5**).

The proportion of abstracts and full texts reporting at least one P-value ≤.05 remained in the 94%-98% range since 1998 (**Figure 3A**). The proportion of reports with at least one P-value ≤.005 demonstrated a gradual increase over time, reaching 57.0% for PubMed abstracts and 62.5% for PMC full texts (**Figure 3B**).

**Figure 3.**
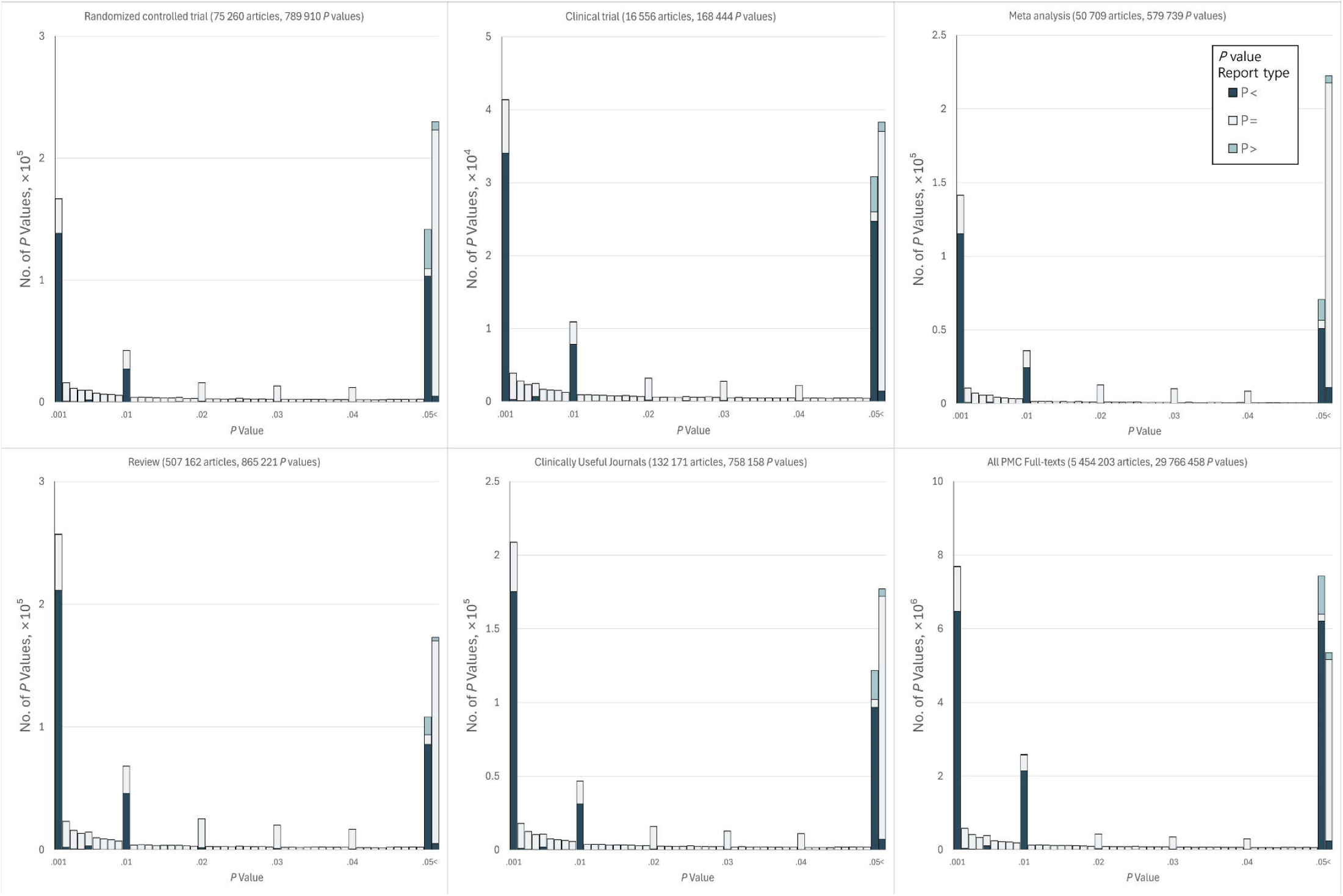
(A) Proportion of articles that have at least 1 *P*-value of .05 or less. (B) Proportion of articles that have at least 1 *P*-value of .005 or less. The proportion is calculated within the subset of all articles that have at least 1 reported *P*-value in PubMed Central abstracts, PubMed Central full-texts, and PubMed abstracts, 1990– 2025. The proportions derived from PMC Full-texts data for the 1990–1996 interval may lack statistical stability, as this is the only period where the number of articles reporting at least one P-value is below 50.

The analysis of P-value report types (classified as ‘<’, ‘=’, and ‘>’) revealed a steady increase in the reporting of ‘exact’ P-values until 2015, but no further substantial increase in the last 10 years (PubMed abstracts 17.6% in 1990, 51.1% in 2015, 49.8% in 2025; PMC abstracts 39.1%, 50.6%, 49.4%, respectively; PMC full-texts 16.7%, 42.3%, 43.4%, respectively) (**Supplementary Figure 6-8**).

Mean P-value counts per article increased between 2000 and 2025, from 2.7 to 3.4 for PubMed abstracts, 2.5 to 3.4 for PMC abstracts, and 3.9 to 12.2 for PMC full-texts. Median counts followed this rising trend (**Supplementary Figure 9-11**).

## Discussion

Despite pleas for moving away from P-values, our evaluation shows that reporting of P-values remains common in abstracts and has become even more common in full text papers. Furthermore, there is an increase in median number of P-values reported per article. Reporting of exact P-values increased until 2015 but no further increase was seen in the last decade. A high proportion of reported values remains clustered around .05, the traditional threshold for statistical significance and almost all abstracts and full-text papers that use P-values have some that are statistically significant by this threshold. Moreover, almost two-thirds of full-texts and slightly lower proportion of abstracts with P-values report at least one P-value that matches the more stringent .005 threshold.

Our evaluation demonstrates the pervasive entrenchment of P-values in the biomedical literature. Despite heavy debates and major changes in the content of the biomedical literature, statistical significance remains almost ubiquitous and there is even a higher volume of reported P-values.^5^ Users and readers of the literature should remain aware about the major issues surrounding P-value misuse and misinterpretation.^1^ The ubiquity of reporting statistically significant P-values extends also beyond biomedicine.^9^

This study has several limitations. First, our regular expression algorithms may have missed P-value reports with non-standard formatting, which could introduce measurement error. However, validation of the algorithms had previously shown low error rates.^7^ Second, while comprehensive, the PubMed and PubMed Central databases do not encompass the entire universe of biomedical and life science literature. Third, our extraction engines generally do not capture P-values in tables, figures, and supplements. However, P-values in abstracts and texts are likely to be the most influential and notable. Another empirical evaluation of tables and figures of some prestigious journals has shown also pervasive presence of statistical significance.^10^ Fourth, we focused on reported P-values without addressing the presence or correction for any multiplicity. It is plausible that for several fields that have moved to more massive data collection and analyses, multiplicity issues may have become more common. However, perhaps there is also more sensitization for handling multiplicity properly. These evolutions may affect the proportion of reported P-values that are below various thresholds.

Finally, we did not examine reporting of effect sizes and confidence intervals. There is evidence that reporting of effect sizes and confidence intervals has improved over time in some study types and journals.^11–13^ However, other evaluations suggest that the vast majority of reported effect sizes in abstracts are also statistically significant anyhow based on concomitantly presented P-values (85% in 2010-2015).^14^ Additional science-wide evaluations may examine how patterns of effect size and uncertainty measures evolve; and whether/how they correlate with potential concomitant use of P-values.

## Data Availability

All data produced in the present study are available upon reasonable request to the authors
All codes produced are available online at github

https://github.com/mizoba/20250923PubMedStudy.git

## Acknowledgement

This work was supported by the Yonsei Fellowship, funded by Lee Youn Jae (JIS).

The work of John Ioannidis is supported by grant N000142412687 from the Office of Naval Research.

