## Supplementary for "Evolution of Reporting P-values Across the Biomedical Literature, 1990-2025: an Updated Meta-Research Study"

**Supplementary Content**

Supplementary Methods

Supplementary Results

Supplementary Table 1. Distribution of the different P-value operators in PubMed abstracts

Supplementary Table 2. Distribution of the classified P-value operators in PubMed abstracts

Supplementary Table 3. Count of extremely small P-values by year.

Supplementary Figure 1. Distribution of P-values in 5,157,951 PMC Abstracts in the Period 2015-2025

Supplementary Figure 2. Distribution of P-values in 10,427,678 PubMed Abstracts in the Period 2015-2025

Supplementary Figure 3. (A) Evolution of the mean of the -log_10_ “best” P-values in PubMed abstracts, 1990-2025

Supplementary Figure 3. (B) Evolution of the mean of the -log_10_ “worst” P-values in PubMed abstracts, 1990-2025

Supplementary Figure 4. (A) Evolution of the mean of the -log_10_ “best” P-values in PMC abstracts, 1990-2025

Supplementary Figure 4. (B) Evolution of the mean of the -log_10_ “worst” P-values in PMC abstracts, 1990-2025

Supplementary Figure 5. (A) Evolution of the mean of the -log_10_ “best” P-values in PMC full-texts, 1990-2025

Supplementary Figure 5. (B) Evolution of the mean of the -log_10_ “worst” P-values in PMC full-texts, 1990-2025

Supplementary Figure 6. Proportion of P-value report type in the period 1990-2025, from All PubMed abstracts

Supplementary Figure 7. Proportion of P-value report type in the period 1990-2025, from All PMC abstracts

Supplementary Figure 8. Proportion of P-value report type in the period 1990-2025, from All PMC full-texts

Supplementary Figure 9. Median of Reported P-value counts per article by year

Supplementary Figure 10. Mean of Reported P-value counts per article by year

Supplementary Figure 11. Trimmed Mean of Reported P-value counts per article by year

Data sharing

References

This supplement provides detailed descriptions of the research methodology and expanded results.

**Supplementary Methods**

We performed automated text-mining analysis on the entire PubMed and PubMed Central (PMC), in the period 1990-2025. For the PubMed analysis, we extracted all P-value data from MEDLINE-indexed articles that have abstracts published between January 1, 1990, and September 23, 2025. The same methodology was applied to the PMC Open Access subset (downloaded October 11, 2025), analyzing both abstracts and the full-texts. We downloaded .xml files for each article in PMC Open Access subset; these files generally do not include figures, tables, and supplements.

**Data Source and Acquisition**

1. PubMed Abstract Acquisition

We performed a systematic extraction of abstracts from the PubMed database using the Entrez Programming Utilities (E-utilities). To ensure data quality and the presence of extractable text, the search query specifically targeted records containing abstracts and categorized under the MEDLINE subset. Data retrieval was executed using a shell script that iteratively queried each year within the defined period. For each year, unique PubMed Identifiers (PMIDs) were fetched via esearch. Subsequently, bibliographic data and abstracts were downloaded in XML format using efetch in batches of 1,000 to optimize throughput and comply with NCBI server guidelines. The relevant fields, specifically the PMID and abstract, were parsed and flattened into a structured text format using the xtract tool from the E-utilities suite.

2. PMC Abstract and Full-text Acquisition

Abstracts and Full-texts were obtained from the PMC Open Access subset. We utilized the NCBI FTP server (ftp://ftp.ncbi.nlm.nih.gov/pub/pmc/oa_bulk/) to perform a bulk download of the dataset. The acquisition was performed using wget in mirroring mode, maintaining the directory structure while excluding index files. The dataset comprised articles provided in XML format, covering the period from 1990 to late 2025. These XML files provided comprehensive access to the main body text.

**Definitions of ‘Abstracts’ and ‘Full-texts’**

We parsed the content based on the structural tags within the extracted XML files to ensure data integrity. For PubMed entries, we utilized the NCBI Entrez Direct utility to fetch and extract content specifically contained within the <AbstractText> element.

For PMC articles, we utilized the PMC Open Access Bulk Data Transfer Service (oa_bulk). Although this service provides both .txt and .xml formats for each article, we exclusively processed the .xml files. The abstract sections of PMC articles were extracted by parsing the content within <abstract> tags. In the context of this study, the term "full-text" specifically refers to the main body of the manuscript. This was extracted by parsing the content enclosed within the <body> tags of the XML files.

**Rationale for XML Parsing over Plain Text (.txt)**

While the previous study has utilized the .txt format for full-text analysis, we deliberately chose the XML format to minimize the risk of misinterpretation.^1^ The .txt files provided in the oa_bulk set contain the entire unstructured content of an article, including metadata (e.g., author names, affiliations, ORCIDs, publication dates) and reference lists. Mining these unstructured files introduces a high probability of false positives (e.g., detecting P-values in reference titles as results). In contrast, parsing the XML structure allows for the precise isolation of the <body> text, ensuring that the analysis is restricted to the narrative of the study.

We acknowledge that relying strictly on XML structure introduces a limitation regarding coverage. Older articles (particularly those from the 1990s) are less likely to contain properly formatted <body> tags in their XML versions compared to their plain text counterparts. Consequently, our method retrieves fewer "full-texts" for earlier years compared to the previous study using .txt files.^1^ However, for recent literature (2020–present), most of the articles are well organized in .xml format that provides accurate parsing and clean information, so we considered this is more suitable format for our analysis.

**Validation of Data Integrity**

We observed intermittent discrepancies during the PubMed abstract retrieval process, where the count of downloaded abstract texts did not always align with the corresponding list of PMIDs or the initial search result counts.

To address this, we developed supplementary validation scripts for PubMed that verify the integrity of the downloaded dataset and automatically retrieve any missing abstracts to ensure completeness.

We also included an integrity check for the PMC Open Access bulk files. Compared to PubMed, the PMC bulk downloader was more stable, with the validation process showing no missing files or errors.

**Search strings for each category in PubMed abstracts**

Consistent with the previous study, we separately examined specific article categories with high clinical relevance.^1^ These categories, defined by MEDLINE indexing, included randomized controlled trial, clinical trial, review, and meta-analysis. While the previous study used the ‘Core Clinical Journals’ category from National Library of Medicine (NLM), the filter 'Core Clinical Journals' was retired in 2021. To maintain consistency with the previous study, we adopted the 'Clinically Useful Journals' list as a successor to the NLM 'Core Clinical Journals' filter.^2^ To prevent data overlap, the 'clinical trial' category excluded articles classified as ‘randomized controlled trial’, and the 'review' category excluded those classified as ‘meta-analysis’, as done previously.^1^

Search strings for each category are as follows:

Randomized controlled trial: "Randomized Controlled Trial"[PT] AND "'$year'"[PDAT] AND hasabstract AND medline[sb]

Clinical trial: "Clinical Trial"[PT] NOT "Randomized Controlled Trial"[PT] AND "'$year'"[PDAT] AND hasabstract AND medline[sb]

Meta-analysis: "Meta-analysis"[PT] AND "'$year'"[PDAT] AND hasabstract AND medline[sb]

Review: "Review"[PT] NOT "Meta-analysis"[PT] AND "'$year'"[PDAT] AND hasabstract AND medline[sb]

Clinically Useful Journals: ("AACN Adv Crit Care"[Journal] OR "Acad Emerg Med"[Journal] OR "Acad Med"[Journal] OR "Addict Behav"[Journal] OR "Age Ageing"[Journal] OR "AJR Am J Roentgenol"[Journal] OR "Allergy"[Journal] OR "Am Fam Physician"[Journal] OR "Am Heart J"[Journal] OR "Am J Epidemiol"[Journal] OR "Am J Gastroenterol"[Journal] OR "Am J Hematol"[Journal] OR "Am J Kidney Dis"[Journal] OR "Am J Med Genet A"[Journal] OR "Am J Med"[Journal] OR "Am J Nurs"[Journal] OR "Am J Obstet Gynecol"[Journal] OR "Am J Prev Med"[Journal] OR "Am J Psychiatry"[Journal] OR "Am J Respir Crit Care Med"[Journal] OR "Am J Sports Med"[Journal] OR "Am J Surg Pathol"[Journal] OR "Am J Med Sci"[Journal] OR "Anesth Analg"[Journal] OR "Ann Allergy Asthma Immunol"[Journal] OR "Ann Emerg Med"[Journal] OR "Ann Intern Med"[Journal] OR "Ann Neurol"[Journal] OR "Ann Oncol"[Journal] OR "Ann Pharmacother"[Journal] OR "Ann Surg"[Journal] OR "Ann Surg Oncol"[Journal] OR "Ann Rheum Dis"[Journal] OR "Ann Thorac Surg"[Journal] OR "Arch Dis Child"[Journal] OR "Arch Dis Child Fetal Neonatal Ed"[Journal] OR "Arch Phys Med Rehabil"[Journal] OR "Arthritis Rheumatol"[Journal] OR "Arthritis Care Res (Hoboken)"[Journal] OR "Arthroscopy"[Journal] OR "Autoimmun Rev"[Journal] OR "Best Pract Res Clin Rheumatol"[Journal] OR "Biol Psychiatry"[Journal] OR "BJOG"[Journal] OR "BJU Int"[Journal] OR "Blood"[Journal] OR "BMJ"[Journal] OR "Bone Marrow Transplant"[Journal] OR "Brain"[Journal] OR "Breastfeed Med"[Journal] OR "Br J Anaesth"[Journal] OR "Br J Cancer"[Journal] OR "Br J Dermatol"[Journal] OR "Br J Haematol"[Journal] OR "Br J Ophthalmol"[Journal] OR "Br Med Bull"[Journal] OR "CA Cancer J Clin"[Journal] OR "Cancer"[Journal] OR "Cancer Treat Rev"[Journal] OR "Catheter Cardiovasc Interv"[Journal] OR "Chest"[Journal] OR "Circulation"[Journal] OR "Clin Biochem"[Journal] OR "Clin Biomech (Bristol)"[Journal] OR "Clin Gastroenterol Hepatol"[Journal] OR "Clin Infect Dis"[Journal] OR "Clin Obstet Gynecol"[Journal] OR "Clin Pharmacol Ther"[Journal] OR "Clin Ther"[Journal] OR "Clin Podiatr Med Surg"[Journal] OR "CMAJ"[Journal] OR "Comput Inform Nurs"[Journal] OR "Crit Care Med"[Journal] OR "Curr Opin Cardiol"[Journal] OR "Curr Opin Gastroenterol"[Journal] OR "Curr Opin Nephrol Hypertens"[Journal] OR "Curr Opin Pediatr"[Journal] OR "Curr Opin Rheumatol"[Journal] OR "Diabetes Care"[Journal] OR "Diabetes Res Clin Pract"[Journal] OR "Diagn Microbiol Infect Dis"[Journal] OR "Dig Dis Sci"[Journal] OR "Dis Colon Rectum"[Journal] OR "Drug Alcohol Depend"[Journal] OR "Early Hum Dev"[Journal] OR "Epilepsia"[Journal] OR "Epilepsy Behav"[Journal] OR "Europace"[Journal] OR "Eur Heart J"[Journal] OR "Eur J Cancer"[Journal] OR "Eur J Cardiothorac Surg"[Journal] OR "Eur J Heart Fail"[Journal] OR "Eur J Intern Med"[Journal] OR "Eur J Nucl Med Mol Imaging"[Journal] OR "Eur J Radiol"[Journal] OR "Eur Urol"[Journal] OR "Fertil Steril"[Journal] OR "Gastroenterology"[Journal] OR "Gastrointest Endosc"[Journal] OR "Gut"[Journal] OR "Gynecol Oncol"[Journal] OR "Head Neck"[Journal] OR "Headache"[Journal] OR "Health Aff (Millwood)"[Journal] OR "Heart"[Journal] OR "Heart Rhythm"[Journal] OR "Hepatology"[Journal] OR "Hum Pathol"[Journal] OR "Hum Reprod"[Journal] OR "Hypertension"[Journal] OR "Infect Control Hosp Epidemiol"[Journal] OR "Int J Antimicrob Agents"[Journal] OR "Int J Cancer"[Journal] OR "Int J Cardiol"[Journal] OR "Int J Clin Pract"[Journal] OR "Int J Obes"[Journal] OR "Int J Radiat Oncol Biol Phys"[Journal] OR "Int Urogynecol J"[Journal] OR "JAMA"[Journal] OR "JAMA Dermatol"[Journal] OR "JAMA Intern Med"[Journal] OR "JAMA Neurol"[Journal] OR "JAMA Ophthalmol"[Journal] OR "JAMA Otolaryngol Head Neck Surg"[Journal] OR "JAMA Pediatr"[Journal] OR "JAMA Psychiatry"[Journal] OR "JAMA Surg"[Journal] OR "J Healthc Qual"[Journal] OR "J Acquir Immune Defic Syndr"[Journal] OR "J Adv Nurs"[Journal] OR "J Allergy Clin Immunol"[Journal] OR "J Altern Complement Med"[Journal] OR "J Bone Joint Surg Am"[Journal] OR "J Card Fail"[Journal] OR "J Clin Endocrinol Metab"[Journal] OR "J Clin Gastroenterol"[Journal] OR "J Clin Neurosci"[Journal] OR "J Clin Oncol"[Journal] OR "J Clin Pathol"[Journal] OR "J Clin Psychol"[Journal] OR "J Clin Psychopharmacol"[Journal] OR "J Emerg Med"[Journal] OR "J Foot Ankle Surg"[Journal] OR "J Gen Intern Med"[Journal] OR "J Hand Surg Am"[Journal] OR "J Hepatol"[Journal] OR "J Hosp Infect"[Journal] OR "J Hosp Med"[Journal] OR "J Infect"[Journal] OR "J Infect Dis"[Journal] OR "J Intern Med"[Journal] OR "J Invest Dermatol"[Journal] OR "J Med Genet"[Journal] OR "J Midwifery Womens Health"[Journal] OR "J Neurol Neurosurg Psychiatry"[Journal] OR "J Nurs Adm"[Journal] OR "J Obstet Gynecol Neonatal Nurs"[Journal] OR "J Occup Environ Med"[Journal] OR "J Oral Maxillofac Surg"[Journal] OR "J Orthop Sports Phys Ther"[Journal] OR "J Orthop Trauma"[Journal] OR "J Pain Symptom Manage"[Journal] OR "J Palliat Med"[Journal] OR "J Pediatr Gastroenterol Nutr"[Journal] OR "J Pediatr Hematol Oncol"[Journal] OR "J Pediatr Orthop"[Journal] OR "J Pediatr Surg"[Journal] OR "J Perinatol"[Journal] OR "J Psychopharmacol"[Journal] OR "J Subst Abuse Treat"[Journal] OR "J Surg Oncol"[Journal] OR "J Am Coll Cardiol"[Journal] OR "J Am Geriatr Soc"[Journal] OR "J Am Med Dir Assoc"[Journal] OR "J Am Med Inform Assoc"[Journal] OR "J Natl Cancer Inst"[Journal] OR "J Thorac Cardiovasc Surg"[Journal] OR "J Thromb Haemost"[Journal] OR "J Trauma Acute Care Surg"[Journal] OR "J Urol"[Journal] OR "J Vasc Surg"[Journal] OR "JPEN J Parenter Enteral Nutr"[Journal] OR "Kidney Int"[Journal] OR "Lancet"[Journal] OR "Laryngoscope"[Journal] OR "Leukemia"[Journal] OR "Liver Transpl"[Journal] OR "Med Care"[Journal] OR "Med Clin North Am"[Journal] OR "Med Lett Drugs Ther"[Journal] OR "Medicine (Baltimore)"[Journal] OR "Mod Pathol"[Journal] OR "Mol Genet Metab"[Journal] OR "Mov Disord"[Journal] OR "Muscle Nerve"[Journal] OR "Nephrol Dial Transplant"[Journal] OR "Neurology"[Journal] OR "Neurosurgery"[Journal] OR "N Engl J Med"[Journal] OR "Nursing"[Journal] OR "Obesity (Silver Spring)"[Journal] OR "Obstet Gynecol Surv"[Journal] OR "Obstet Gynecol"[Journal] OR "Oral Surg Oral Med Oral Pathol Oral Radiol"[Journal] OR "Otolaryngol Head Neck Surg"[Journal] OR "Pain"[Journal] OR "Pain Med"[Journal] OR "Patient Educ Couns"[Journal] OR "Pediatr Dermatol"[Journal] OR "Pediatr Infect Dis J"[Journal] OR "Pediatrics"[Journal] OR "Pharmacoepidemiol Drug Saf"[Journal] OR "Plast Reconstr Surg"[Journal] OR "Postgrad Med J"[Journal] OR "Prev Med"[Journal] OR "Prim Care"[Journal] OR "Psychiatr Serv"[Journal] OR "QJM"[Journal] OR "Radiographics"[Journal] OR "Radiology"[Journal] OR "Radiother Oncol"[Journal] OR "Respir Med"[Journal] OR "Semin Dial"[Journal] OR "Semin Nucl Med"[Journal] OR "Semin Perinatol"[Journal] OR "Semin Respir Crit Care Med"[Journal] OR "Semin Ultrasound CT MR"[Journal] OR "Sex Transm Dis"[Journal] OR "Sex Transm Infect"[Journal] OR "Soc Sci Med"[Journal] OR "South Med J"[Journal] OR "Spine (Phila Pa 1976)"[Journal] OR "Stat Methods Med Res"[Journal] OR "Stat Med"[Journal] OR "Stroke"[Journal] OR "Thorax"[Journal] OR "Thromb Res"[Journal] OR "Thyroid"[Journal] OR "Ultrasound Obstet Gynecol"[Journal] OR "Vaccine"[Journal] OR "World Neurosurg"[Journal]) AND "'$year'"[PDAT] AND hasabstract AND medline[sb]

All PubMed: "'$year'"[PDAT] AND hasabstract AND medline[sb]'

The search queries for the "Clinically Useful Journals" category were derived from the reference.^2^ However, the specific term "Clin Biomech (Bristol, Avon)"[Journal] yielded zero results because the search term of the journal in PubMed (NLM index based) has changed. We identified the updated term as "Clin Biomech (Bristol)"[Journal] and revised the search string accordingly.

**Detection algorithm for P-values**

A 'P-value report' was defined as a text string comprising a variation of the term 'P-value’, followed by a relational operator and a numeric value. The search string for P-values is given below:

p_value_pattern = r"""(?x)	# Verbose mode (ignore whitespace in regex)

(\s|\()	# Preceding space or parenthesis

[Pp]{1}	# 'P' or 'p'

(\s|-)* # Optional space or hyphen

(value|values)? # Optional 'value' word

(\s)* # Optional space

(?P<op>	# START GROUP: Operator

([=<>≤≥]|less\s+than|of\s+<)+	# Matches signs, 'less than', 'of <'S$

)	# END GROUP: Operator

(\s)* # Optional space

(?P<num>	# START GROUP: Number/Value

(?P<digits>([0-9]|([\,\.•][0-9]))	# First digit or separator+digit

[0-9]* # Following digits

[\,\.•]? # Optional separator

[0-9]*) # Decimal digits

(\s)* # Space before suffix

(	# Suffix group (percentage or sci notation)

(\%)|	# Percentage sign

([x×]?(\s)*(?P<base>[0-9]*)(\s)*((exp|Exp|E|e)?(\s)*(?P<exponent>((\((\s)*(-){1}(\s)*[0-9]+(\s)*\))|((\s)*(-){1}(\s)*[0-9]+))))?)

)

)	# END GROUP: Number

"""

The complete source code is available in the data sharing section.

We adopted the P-value detection algorithm and search strings from the previous study, as follows: "/(\s|\()[Pp]{1}(\s|-)*(value| values)?(\s)*([=<>≤≥]|less than|of <)+(\s)*([0-9]|([\,|\.|•][0-9]))[0-9]*[\,|\.|•]?[0-9]*(\s)*((\%)|([x|×]?(\s)*[0-9]*(\s)*((exp|Exp|E|e)?(\s)*((\((\s)*(-){1}(\s)*[0-9]+(\s)*\))|((\s)*(-){1}(\s)*[0-9]+)))?))/".

The original algorithm had been validated via manual extraction, demonstrating high performance (96.3% sensitivity and 99.8% specificity). To maintain consistency with this baseline performance while ensuring stability on newer datasets, we performed one essential syntax correction.

Correction of Regular Expression Syntax

We identified a recurrent syntax issue in the original regular expressions where the pipe symbol (|) was incorrectly used within Character Classes ([ ]) as an intended "OR" operator.

In standard regular expressions, the pipe symbol functions as a logical "OR" only within grouping parentheses (e.g., (A|B)). Inside square brackets, however, it is interpreted as a literal vertical bar character. We systematically removed these redundant pipe symbols from all Character Class definitions in the detection strings (for instance, correcting [\,|\.|•] to [\,\.•], among other**s**).

This correction was critical because the original pattern unintentionally identified vertical bars—often found in table delimiters or mathematical notation—as valid parts of numeric values. A notable consequence was that Python's string-to-float conversion would fail when encountering these bars within potential P-value strings. We suspect this syntax issue remained undetected in the previous study (conducted in 2015) because the specific text patterns triggering these errors were absent in the PubMed abstract dataset before 2015. We have corrected this to ensure robust processing of the full, modern database.

Refinements in the post-processing of P-values

Following extraction, we implemented five specific refinements to the post-processing phase to enhance data accuracy and coverage compared to the original study.

**1. Validity Interval:** We excluded P-values not in the interval (0,1) while the original article excluded P-values not in the interval [0,1], assuming that these were probably not referring to P-values.

**2. Handling of Percentages:** While the original article discarded P-value expressions followed by a ‘%’ sign even though they detected them, we included these P-values and normalized them by multiplying by 0.01.

**3. Classification of Operators:** When we classified the operators detected by the search string, we standardized the wide variety of operator strings found in the literature into three primary categories: “Less than” (<), “Greater than” (>), and “Equal to” (=). “Less than” includes strict inequalities (<, <<) and non-strict inequalities such as ≤, <=, =<. “Greater than” includes strict inequalities (>, >>) and non-strict inequalities such as ≥, >=, =>. “Equal to” includes standard equalities (=, ==), and “Ambiguous” includes strings containing contradictory operators (e.g., ><, <>) and were excluded from the analyses of operator proportions. The full distribution of raw strings and their classification is detailed in **Supplementary Table 1,2**.

**4. Inclusion of "Greater Than" Values:** Unlike the previous study, which excluded P-values with "Greater than" expressions when characteristics of P-values were calculated, we included these values in analysis to monitor the entirety of P-value reporting.

**5. Correction of Exponential Notation:** We implemented a safeguard for exponential notation errors where the base was detected as zero, treating the base as 10. In the P-value search string, the part containing a minus (-) is considered the exponent term, which is “(?P<exponent>((\((\s)*(-){1}(\s)*[0-9]+(\s)*\))|((\s)*(-){1}(\s)*[0-9]+)))”. The part preceding the exponent, consisting of numbers adjacent to ‘x’ or ‘×’ or ‘exp’ or ‘Exp’ or ‘E’ or ‘e’, is considered the base term, in the P-value search string “([x|×]?(\s)*(?P<base>[0-9]*)(\s)*((exp|Exp|E|e)?”. If the search string detected the base as zero, we assumed the P-value had a base 10, maintaining consistency with the assumptions of the previous study. (In fact, in the PubMed abstract database having 22,734,796 total article and 10,697,189 total P-values, only 4 P-values were detected with a base of zero. Each PMID and P-value in the article was: PMID: 10716308, ‘P<5x0.000001’, PMID: 9726033, ‘P = 0 x 0.03’, PMID: 9726034, ‘P < 0 x 0.001’, PMID: 24962670, ‘p = 1.33 × 0(-5)’. ) Given the sample size, the impact of these interpretations on the overall results is negligible.

**Extent of Consistency with the Original Study**

We verified that our refined search string (while more robust against syntax errors) still detected over 99% of the P-values identified by the original search string.

To further demonstrate the validity of our algorithm, we compared our results for the 2014 PubMed abstract dataset against the values reported in the original article. The proportions of abstracts reporting at least one P-value across categories are:

- Randomized Controlled Trial) Original study: 54.8% vs. Current study: 54.0%
- Clinical Trial) Original study: 38.9% vs. Current study: 39.6%
- Meta-analysis) Original study: 35.7% vs. Current study: 38.1%
- Review) Original study: 2.4% vs. Current study: 1.0%
- Core Clinical Journals) Original study: 33.0% vs. Current study: 33.8%

We believe the minor variations observed are due to the dynamic nature of the PubMed database, which frequently updates publication types and article inclusion. (e.g., many articles previously classified as Reviews are now reclassified as Meta-analyses, resulting in a lower proportion of abstracts reporting P-values). For comparison, we utilized the ‘Core Clinical Journals’ filter, although it was retired that same year.

**Main Outcomes**

We evaluated the following characteristics of P-values and their trends over time: proportion of abstracts and full-texts containing P-values **(Figure 1)**; distribution of reported P-values **(Figure 2, Supplementary Figure 1,2)**; minimum (most statistically significant) and maximum (least statistically significant) P-values reported **(Supplementary Figure 3-5)**; proportion of abstracts and full-texts reporting at least one P-value below the thresholds of .05 and .005 **(Figure 3)**; distribution of relational operators used in reported P-values **(Supplementary Figure 6-8)**; and number of P-values per paper **(Supplementary Figure 9-11)**.

Analyses were performed using Python version 3.11.12 (Python Software Foundation) and Microsoft Excel (Microsoft Corp).

**Supplementary Results**

**Proportion of abstracts and full-texts containing P-values**

From 1990 to 2025, PubMed database included 22,734,796 articles with an abstract, of which 3,500,485 abstracts reported P-values (**Figure 1A**). Proportion of abstracts reporting P-values increased annually from 7.5% (95% CI, 7.4%-7.6%) in 1990 to 17.3% (95% CI, 17.2%-17.4%) in 2015 and since then has remained relatively stable with 18.3% (95% CI, 18.2%-18.4%) in 2025). In 2025, the highest proportion was 58.3% (95% CI, 57.5%-59.0%) in randomized controlled trials, followed by 41.0% (95% CI, 40.2%-41.8%) in meta-analyses, 31.7% (95% CI, 31.3%-32.1%) in clinically useful journals, 30.5% (95% CI, 28.8%-32.2%) in clinical trials, and 1.0% (95% CI, 0.9%-1.1%) in reviews. The increase over time pertains to all categories except for reviews and clinically useful journals, which have shown declines starting in 2024 and 2017, respectively.

We also evaluated 6,031,459 PMC abstracts and 6,397,787 PMC full-texts. The proportion in PMC full-texts was 5.2% (95% CI, 2.3%-8.1%) in 1990, 48.9% (95% CI, 48.7%-49.1%) in 2015 and 53.3% (95% CI, 53.2%-53.5%) in 2025 (**Figure 1B**).

**Distribution of reported P-values**

To check the distribution of reported P-values, we created 50 intervals, each with a width of .001, raging from ‘*P*≤.001’ to ‘.049<*P*≤.050’. (e.g. P-value higher than .034 and equal to or smaller than .035 was assigned to to the interval ‘.034<*P*≤.035’ ). The interval ‘.05<*P’* was added to capture P-values higher than .05. While the previous study assessed the distribution of P-values through 2015, we evaluated the subsequent period from 2015 to 2025 to provide a contemporary update. In all databases, the distribution of P-values showed strong clustering around .05 and .001, followed by .01 **(Figure 2, Supplementary Figure 1, 2)**. In PubMed abstracts, 29.6% of reported P-values fell within the ‘P≤.001’ intereval, 16.6% were in ‘.049<*P*≤.050’, and 13.0% were in ‘.05<*P’*. In PMC full-texts, 25.8% were in ‘*P*≤.001’, 25.0% were in ‘.049<*P*≤.050’, and 18.0% were in ‘.05<*P’*. The ratio of ‘.049 <*P*≤.050’ to ‘*P*≤.001’ was 0.97 and the ratio of ‘.05<*P’* to ‘*P*≤.001’ was 0.70 in PMC full-text articles. These ratios were lower in abstracts: 0.59 and 0.40 for PMC abstracts, and 0.56 and 0.44 for PubMed abstracts, respectively. The less-than operator (‘<’) was most prevalent for ‘*P*≤.001’ and ‘.049<*P*≤.050’, and the equal-to operator (‘=’) was most prevalent for ‘.05<*P’*.

**Minimum and maximum P-values reported**

Mean -log_10_ of minimum P-values was largely stagnant or slightly increasing (2.33 in 1990 to 2.45 in 2025) in PubMed abstracts, was steady in PMC full-texts, and slightly decreased in PMC abstracts (2.53 in 1990 to 2.45 in 2025). Mean -log_10_ of maximum P-values decreased from 1.80 in 1990 to 1.57 in 2025 and the decrease was also consistent in PMC databases (**Supplementary Figure 3-5**).

A peak was observed around 2010 in meta-analysis from mean -log_10_ of minimum P-values in PubMed abstracts. Following a check for outliers, we have investigated the raw data. We specifically extracted P-values smaller than 10^-30^ to identify potential driving factors. The counts of these extremely small P-values were relatively high in the period 2010-2014 and declined thereafter **(Supplementary Table 3)**. We confirmed that over 90% of these extreme values (e.g., 1.4x10-269) stemmed from genetic studies, including Genome-Wide Association Studies (GWAS). Of the 83 P-values identified in 2010-2014, only 4 P-values were not from genetic studies (Environmental Toxicology [n=1], Clinical Pharmacology [n=1], and Epidemiology/Proteomics [n=2]).

We hypothesize that this trend reflects the historical trajectory of GWAS research. The period from 2005 to 2015 focused on identifying common genetic variants with large effect sizes in massive populations, generating extremely small P-values.^3^ However, the field has shifted toward sequencing rare variants and utilizing Polygenic Risk Scores (PRS). This trend moved the focus from isolating single variants with extreme significance (e.g., P=10^-100^) to aggregating thousands of marginally significant variants (e.g., P=10^-5^).^4^ Since genetic meta-analyses acted as outliers that inflated the average, it appears the shift in GWAS focus led to the decrease in -log_10_ of minimum P-values.

**Proportion of abstracts and full-texts reporting at least one P-value below the thresholds of .05 and .005**

The proportion of abstracts and full-texts reporting at least one P-value ≤ .05 remained in the [94%-98%] range since 1998; having a slight decrease for PubMed abstracts until 2018 and a slight increase for PMC full-texts until 2012 but no change in more recent years (**Figure 3A**). The proportion of reports with at least one P-value ≤.005 demonstrated a gradual increase over time, reaching 57.0% (95% CI, 56.7%-57.2%) for PubMed abstracts and 62.5% (95% CI, 62.3%-62.7%) for PMC full-texts (**Figure 3B**).

**Distribution of relational operators used in reported P-values**

The analysis of P-value report types (classified as ‘<’, ‘=’, and ‘>’) revealed a steady increase in the reporting of ‘exact’ P-values until 2015, but no further substantial increase in the last 10 years (PubMed abstracts 17.6% in 1990, 51.1% in 2015, 49.8% in 2025; PMC abstracts 39.1%, 50.6%, 49.4%, respectively; PMC full-texts 16.7%, 42.3%, 43.4%, respectively) (**Supplementary Figure 6-8**).

**Number of P-values per paper**

Mean P-value counts per article increased between 2000 and 2025, from 2.7 to 3.4 for PubMed abstracts, 2.5 to 3.4 for PMC abstracts, and 3.9 to 12.2 for PMC full-texts. Median counts and 5% trimmed mean followed this rising trend **(Supplementary Figure 9-11)**.

| Category | Raw String(s) Found | Total Count |
| --- | --- | --- |
| < (Less than) | < | 5038242 |
|  | of< | 4235 |
|  | lessthan | 125472 |
|  | << | 815 |
|  | <<< | 11 |
|  | of<< | 1 |
|  | =lessthan | 4 |
|  | <== | 23 |
|  | =< | 3085 |
|  | =≤ | 20 |
|  | ≤ | 87469 |
|  | <= | 173 |
|  | ≤= | 6 |
|  | of<= | 3 |
|  | ≤< | 1 |
|  | ≤=< | 1 |
| > (Greater than) | > | 302198 |
|  | >> | 57 |
|  | >>> | 1 |
|  | ≥ | 10458 |
|  | >= | 17 |
|  | ≥= | 2 |
|  | => | 45 |
| = (Equal to) | = | 5124815 |
|  | == | 19 |
| Ambiguous | >< | 14 |
|  | <> | 2 |

**Supplementary Table 1. Distribution of the different P-value operators in PubMed abstracts**

| Category | Total Count |
| --- | --- |
| < | 5259561 |
| = | 5124834 |
| > | 312778 |
| Ambiguous | 16 |

**Supplementary Table 2. Distribution of the classified P-value operators in PubMed abstracts**

| Year | Count |
| --- | --- |
| 2005 | 0 |
| 2006 | 0 |
| 2007 | 2 |
| 2008 | 1 |
| 2009 | 5 |
| 2010 | 12 |
| 2011 | 11 |
| 2012 | 17 |
| 2013 | 21 |
| 2014 | 22 |
| 2015 | 9 |
| 2016 | 11 |
| 2017 | 8 |
| 2018 | 4 |
| 2019 | 3 |
| 2020 | 0 |
| 2021 | 0 |
| 2022 | 9 |
| 2023 | 12 |
| 2024 | 5 |
| 2025 | 3 |

**Supplementary Table 3. Count of extremely small P-values by year.** The table shows counts of extremely small P-values ( < 10^-30^ ) detected in the PubMed Abstract database.

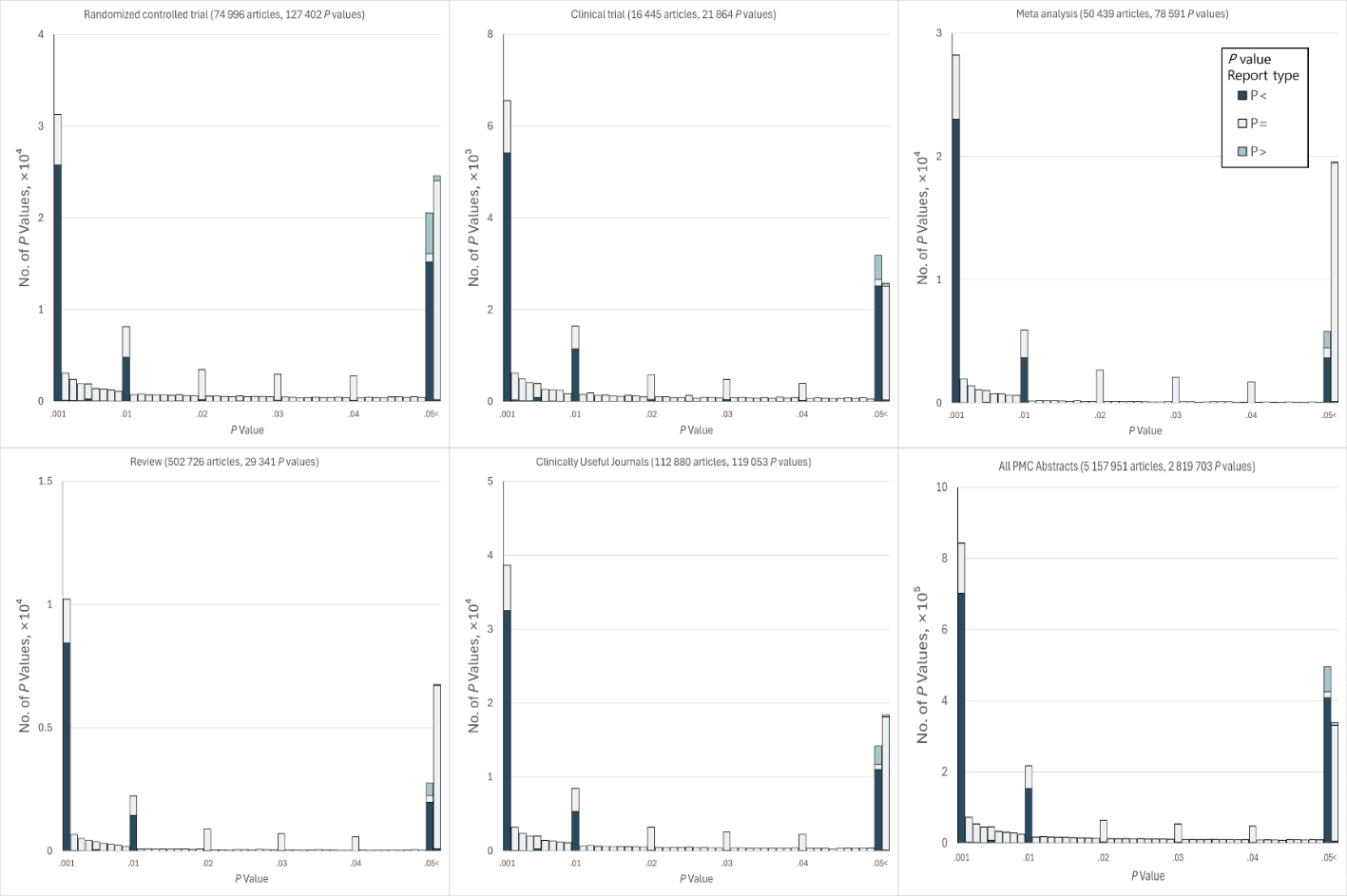

**Supplementary Figure 1. Distribution of P-values in 5 157 951 PMC Abstracts in the Period 2015-2025.** There are total 51 bins shown, each with a width of 0.001, except for the rightmost bin, which represents the number of non-significant *P*-values (0.05<*P*). The categories displayed are: Randomized controlled trial, Clinical trial, Meta-analysis, Review, Clinically Useful Journals, Observational study, Systematic review, and All PubMed Central Abstracts.

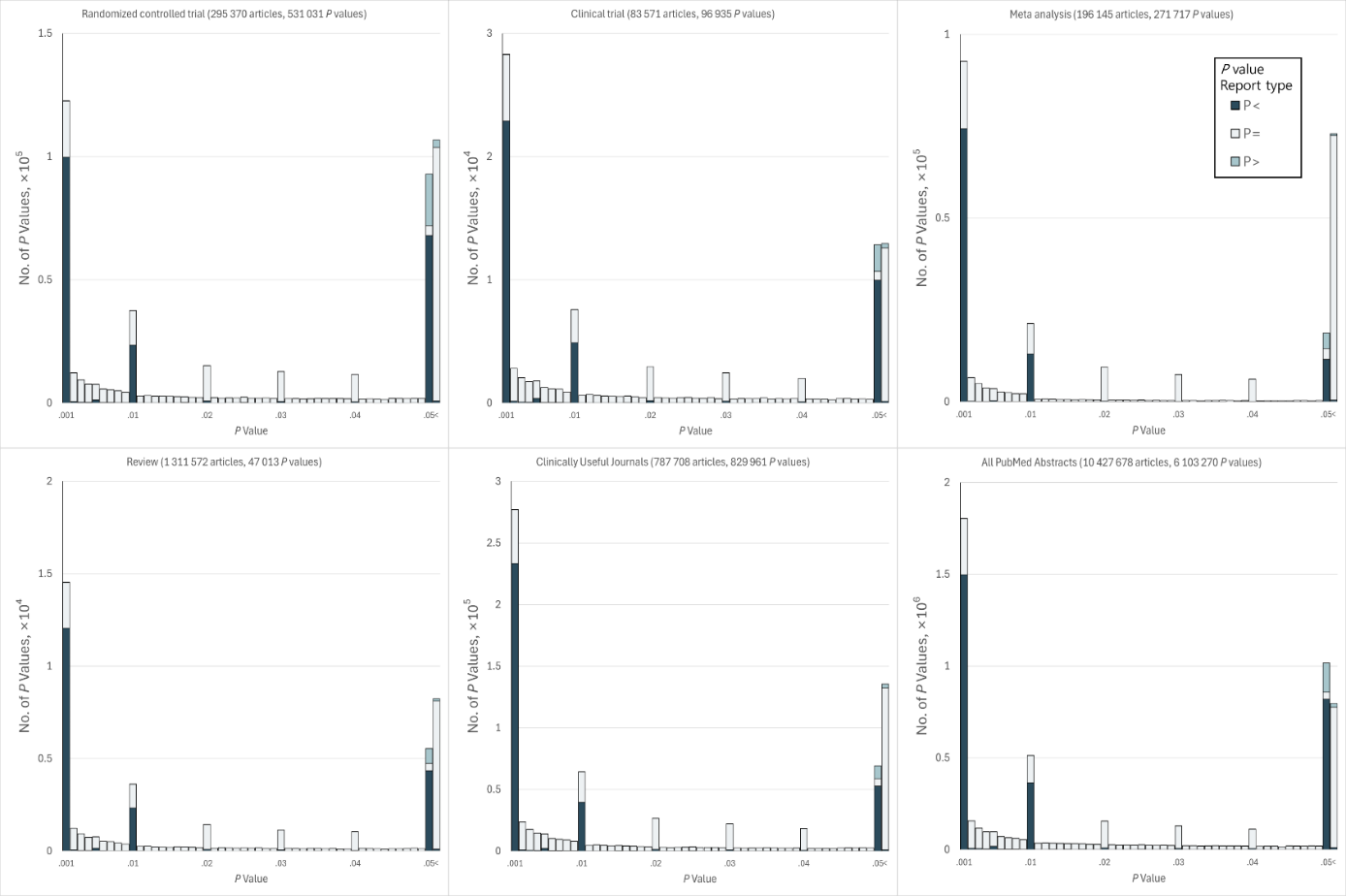

**Supplementary Figure 2. Distribution of P-values in 10 427 678 PubMed Abstracts in the Period 2015-2025.** There are total 51 bins shown, each with a width of 0.001, except for the rightmost bin, which represents the number of non-significant *P*-values (0.05<*P*). The categories displayed are: Randomized controlled trial, Clinical trial, Meta-analysis, Review, Clinically Useful Journals, Observational study, Systematic review, and All PubMed Abstracts.

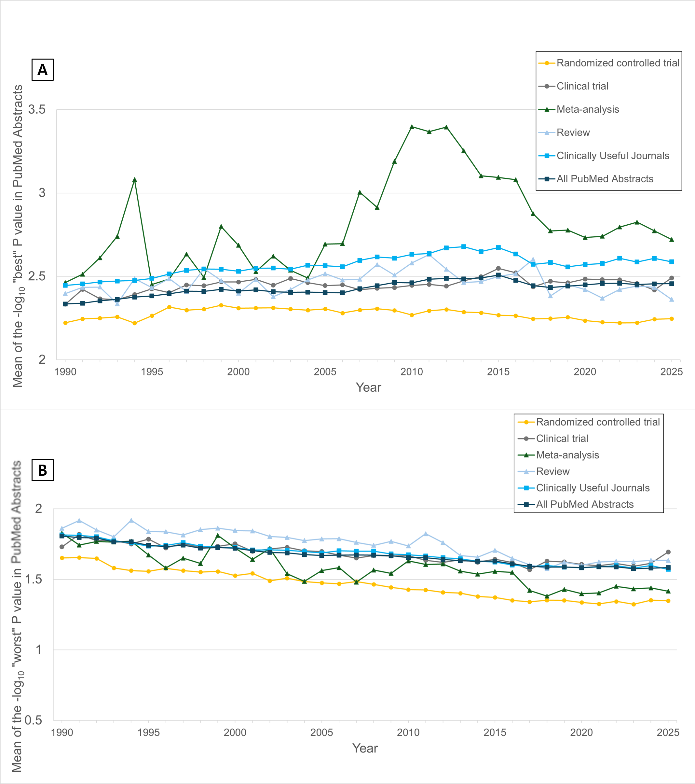

**Supplementary Figure 3. (A) Evolution of the mean of the -log_10_ “best” *P*-values in PubMed abstracts, 1990-2025. (B) Evolution of the mean of the -log_10_ “worst” *P*-values in PubMed abstracts, 1990-2025.** To minimize noise from sparse data for each category, year range continuously maintaining a minimum of N > 5 published articles with at least 1 *P*-value in any given year were retained for inclusion across the entire reporting period.

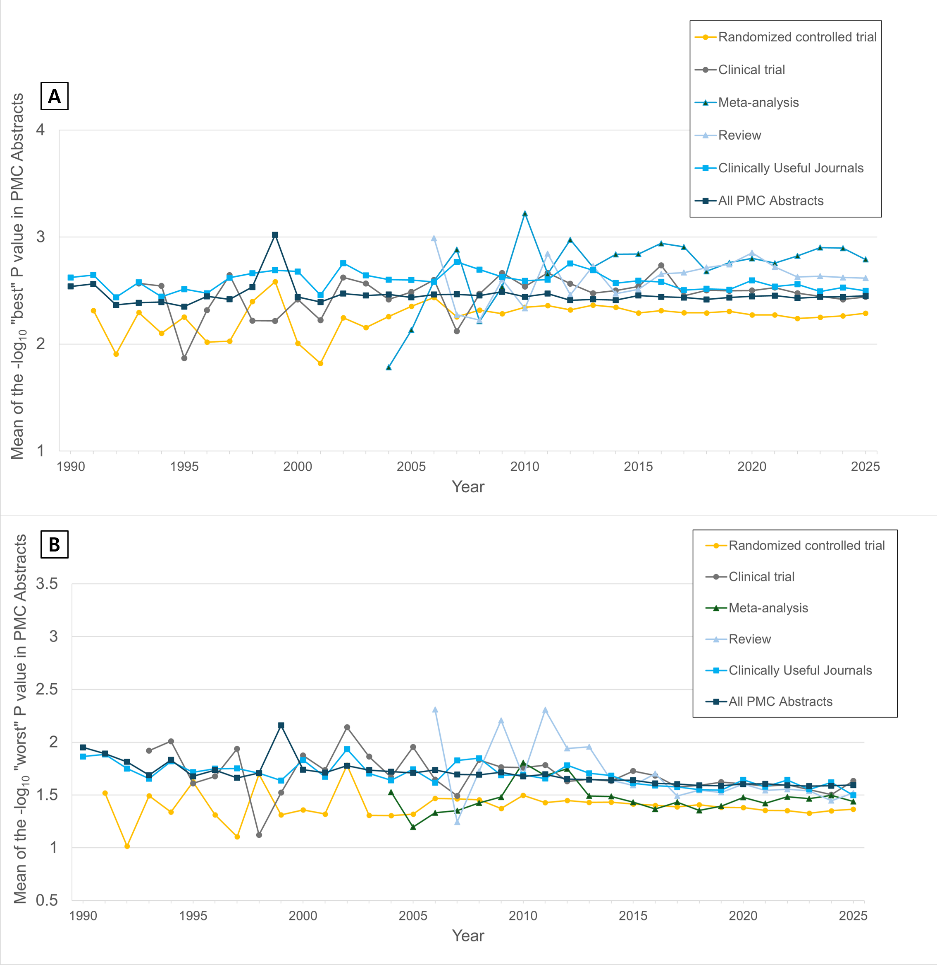

**Supplementary Figure 4. (A) Evolution of the mean of the -log_10_ “best” *P*-values in PMC abstracts, 1990-2025. (B) Evolution of the mean of the -log_10_ “worst” *P*-values in PMC abstracts, 1990-2025.** To minimize noise from sparse data for each category, year range continuously maintaining a minimum of N > 5 published articles with at least 1 *P*-value in any given year were retained for inclusion across the entire reporting period.

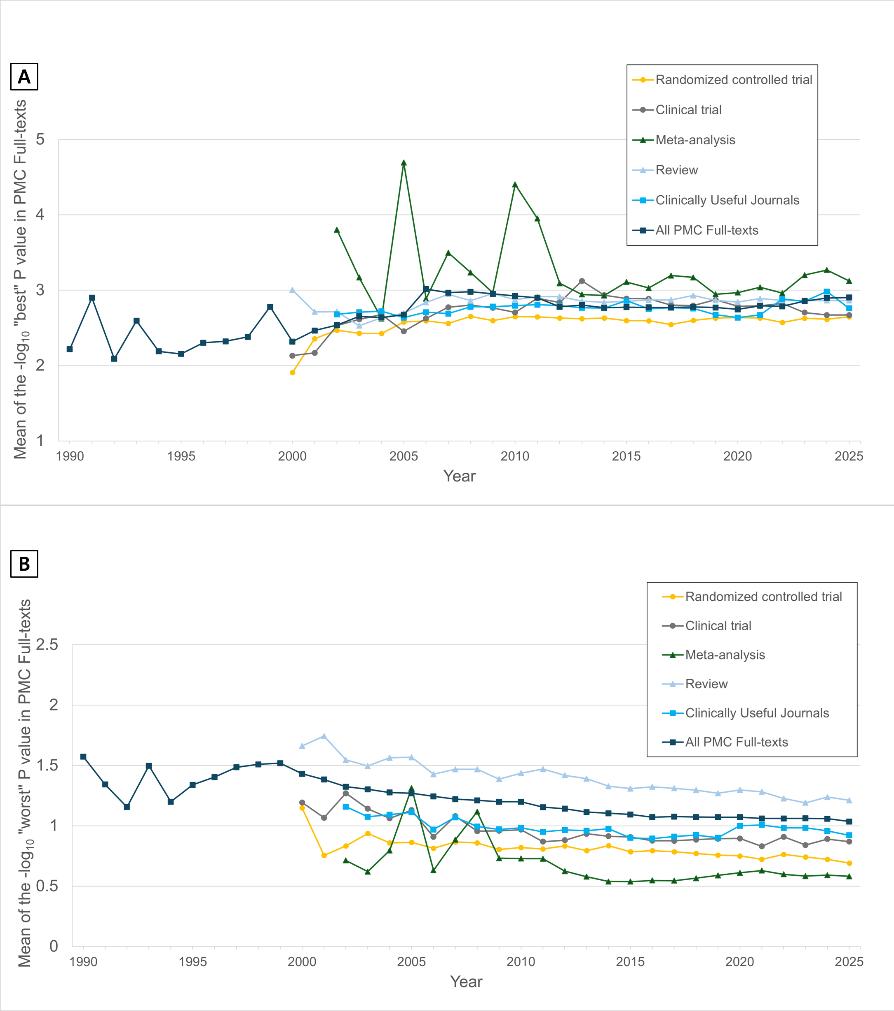

**Supplementary Figure 5. (A) Evolution of the mean of the -log_10_ “best” *P*-values in PMC full-texts, 1990-2025. (B) Evolution of the mean of the -log_10_ “worst” *P*-values in PMC full-texts, 1990-2025.** To minimize noise from sparse data for each category, year range continuously maintaining a minimum of N > 5 published articles with at least 1 *P*-value in any given year were retained for inclusion across the entire reporting period.

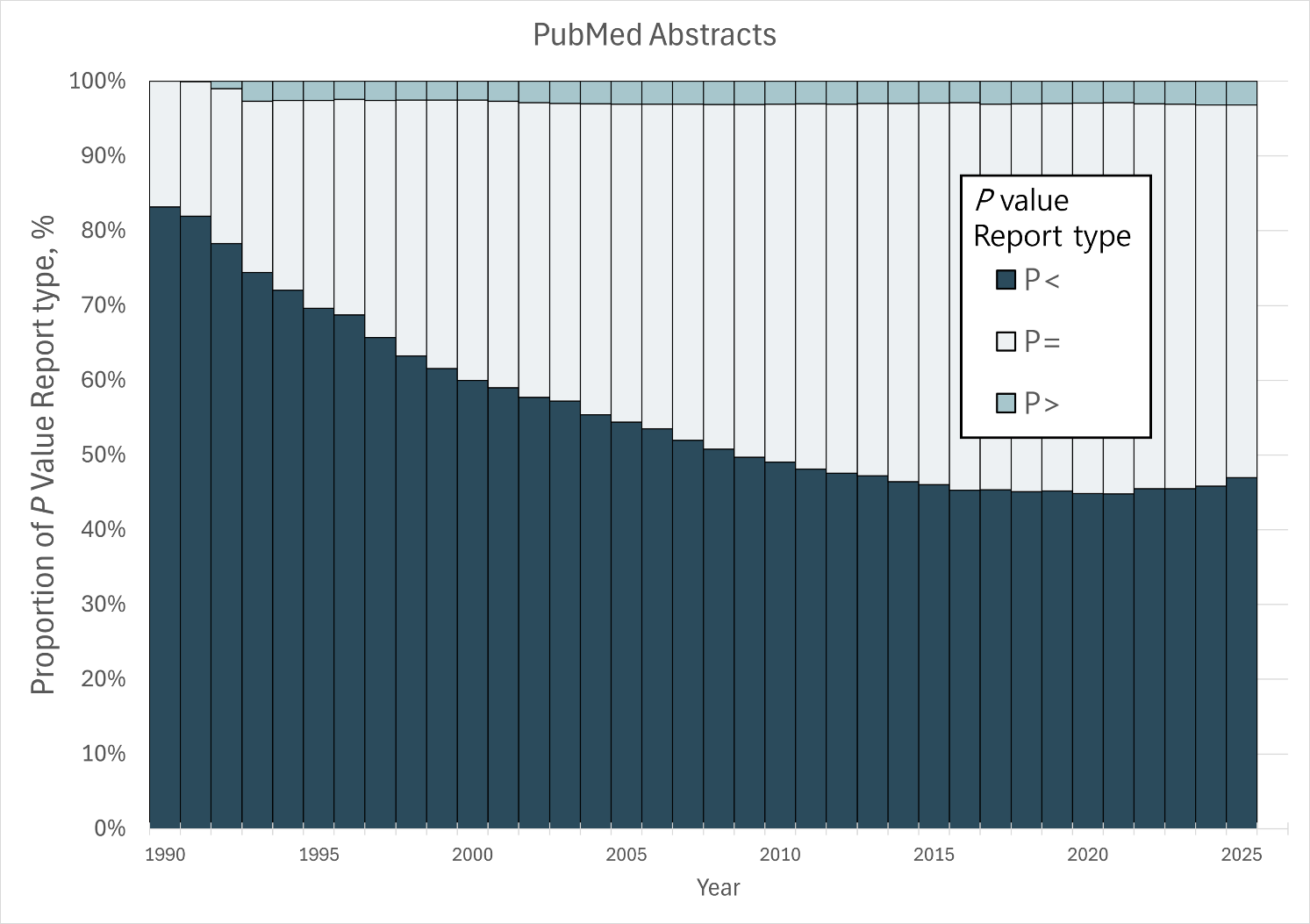

**Supplementary Figure 6. Proportion of *P*-value report type in the period 1990-2025, from All PubMed abstracts.**

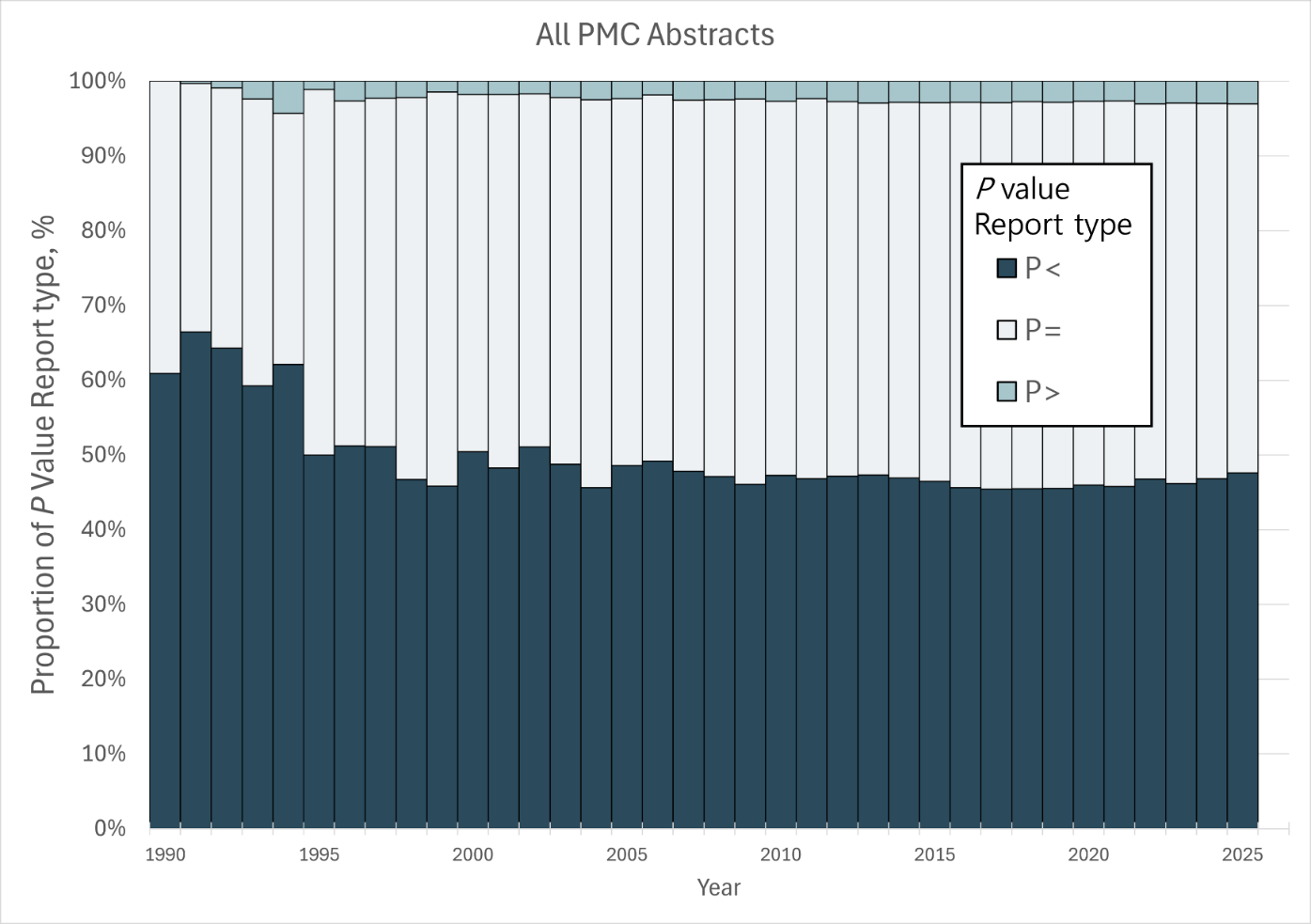

**Supplementary Figure 7. Proportion of *P*-value report type in the period 1990-2025, from All PMC abstracts.**

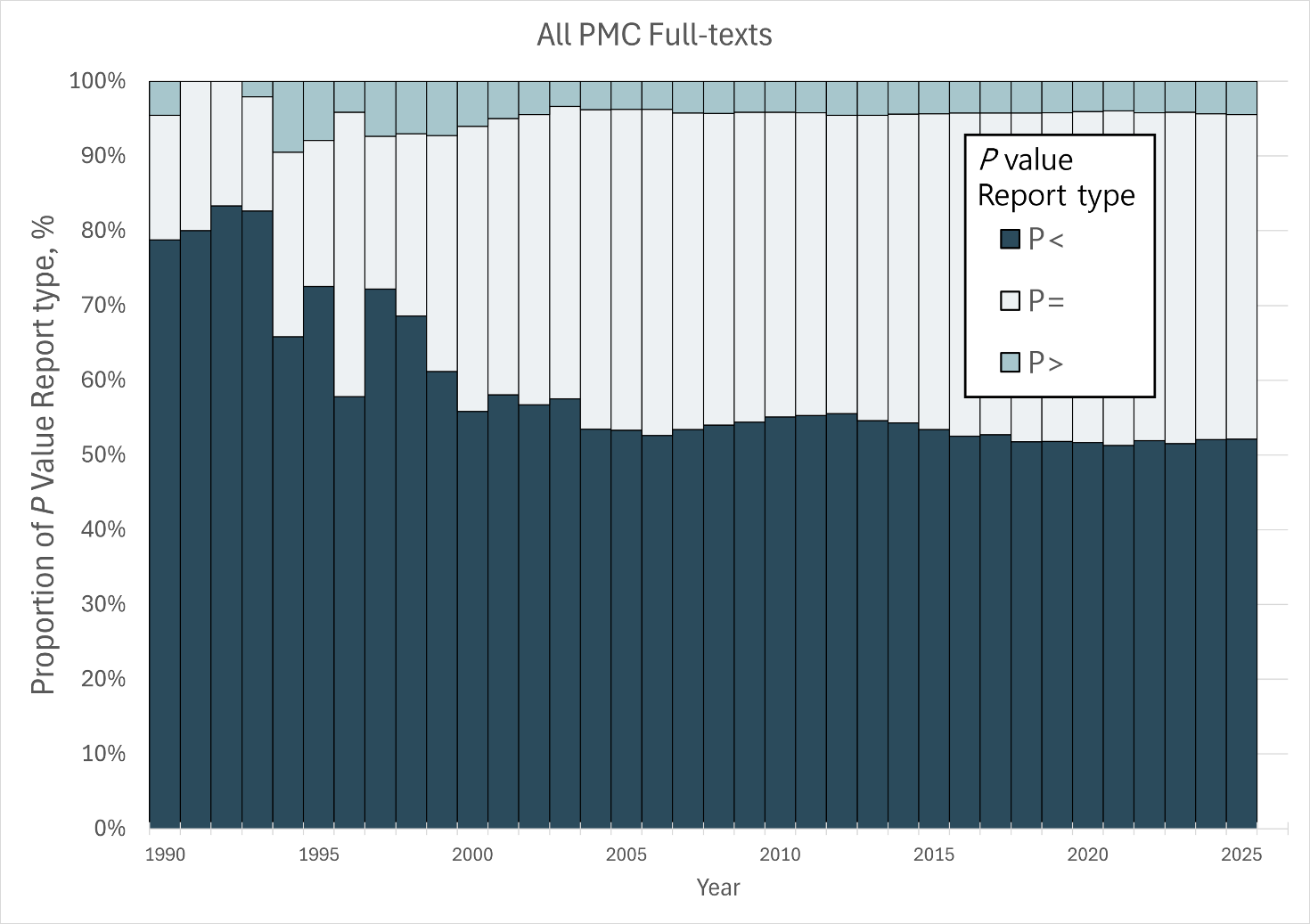

**Supplementary Figure 8. Proportion of *P*-value report type in the period 1990-2025, from All PMC full-texts.**

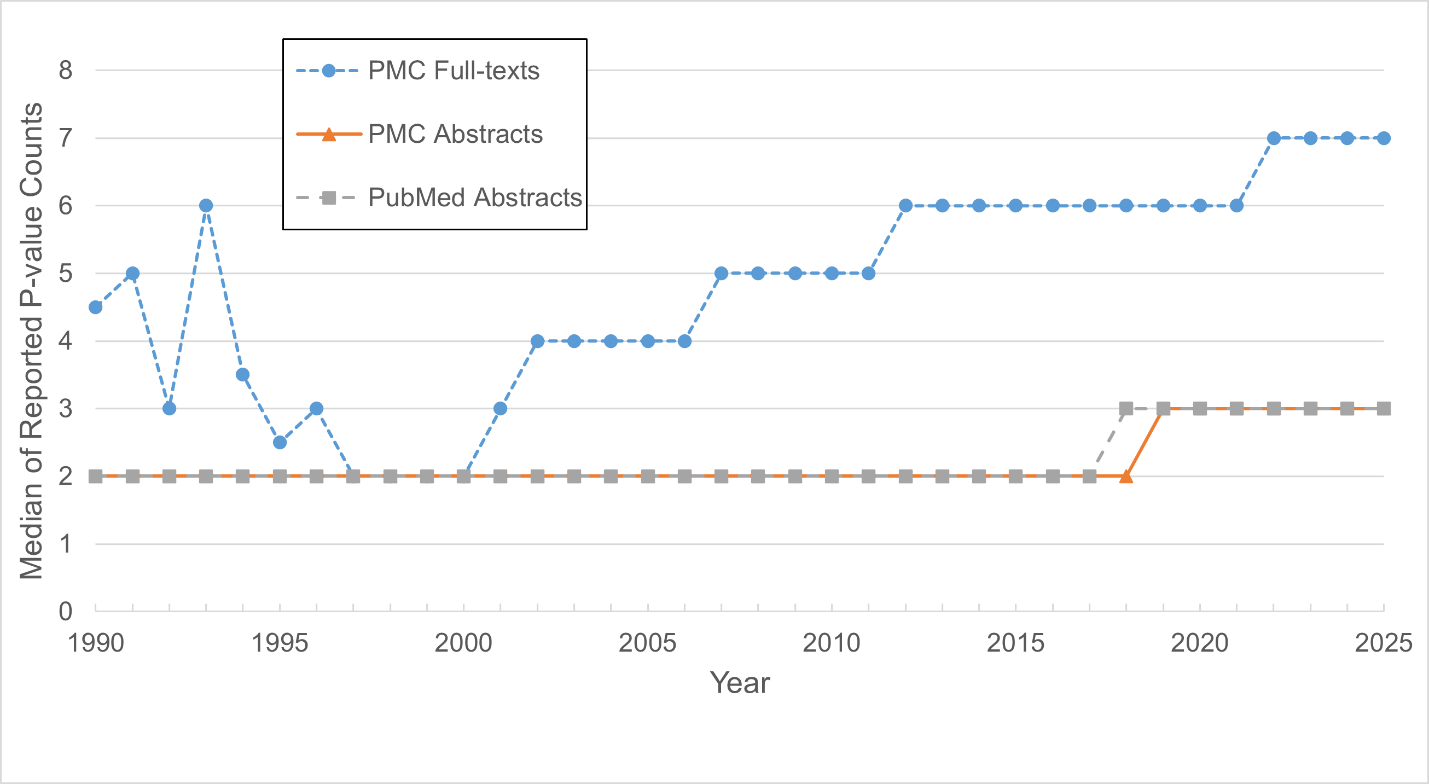

**Supplementary Figure 9. Median of Reported P-value counts per article by year.** The proportions derived from PMC Full-texts data for the 1990–1996 interval may lack statistical stability, as this is the only period where the number of articles reporting at least one P-value is below 50.

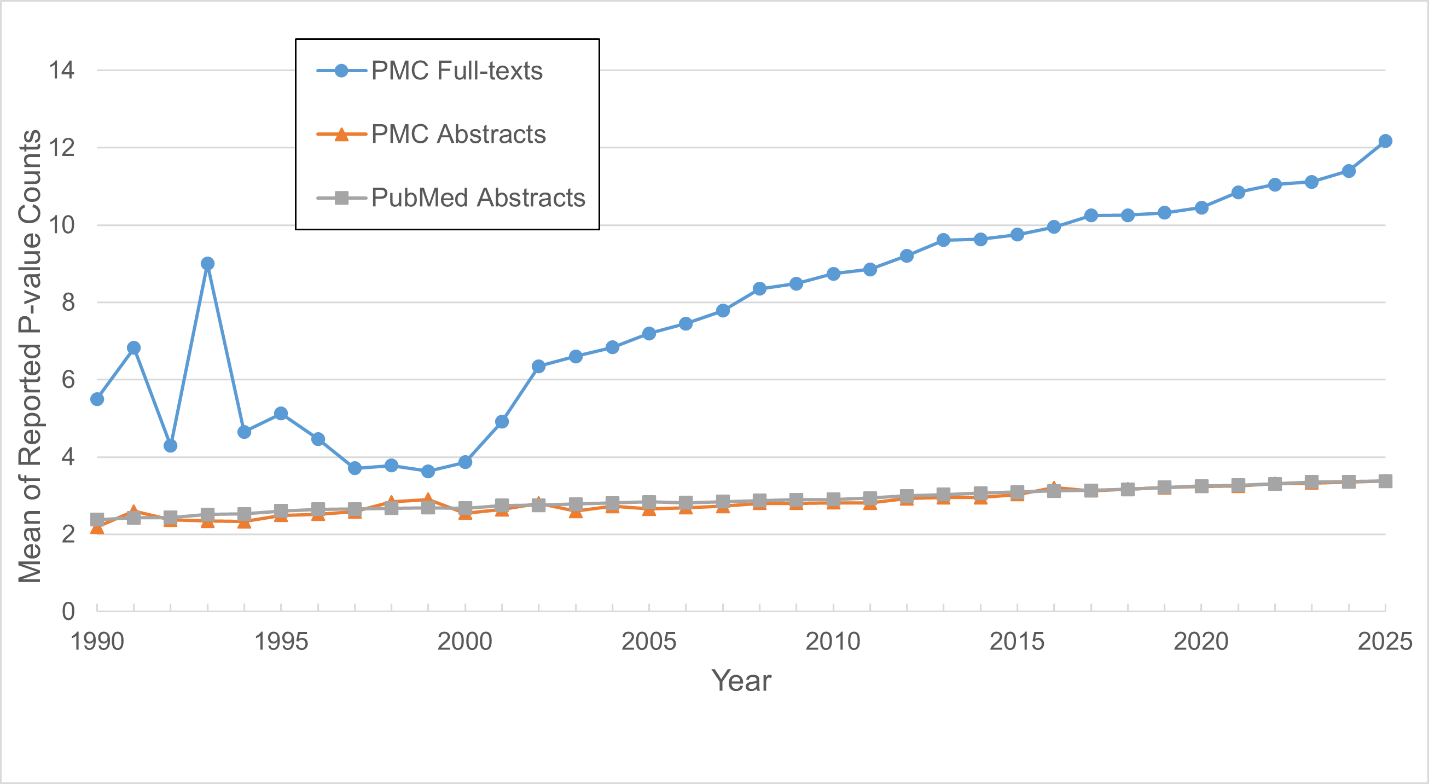

**Supplementary Figure 10. Mean of Reported P-value counts per article by year.** The proportions derived from PMC Full-texts data for the 1990–1996 interval may lack statistical stability, as this is the only period where the number of articles reporting at least one P-value is below 50.

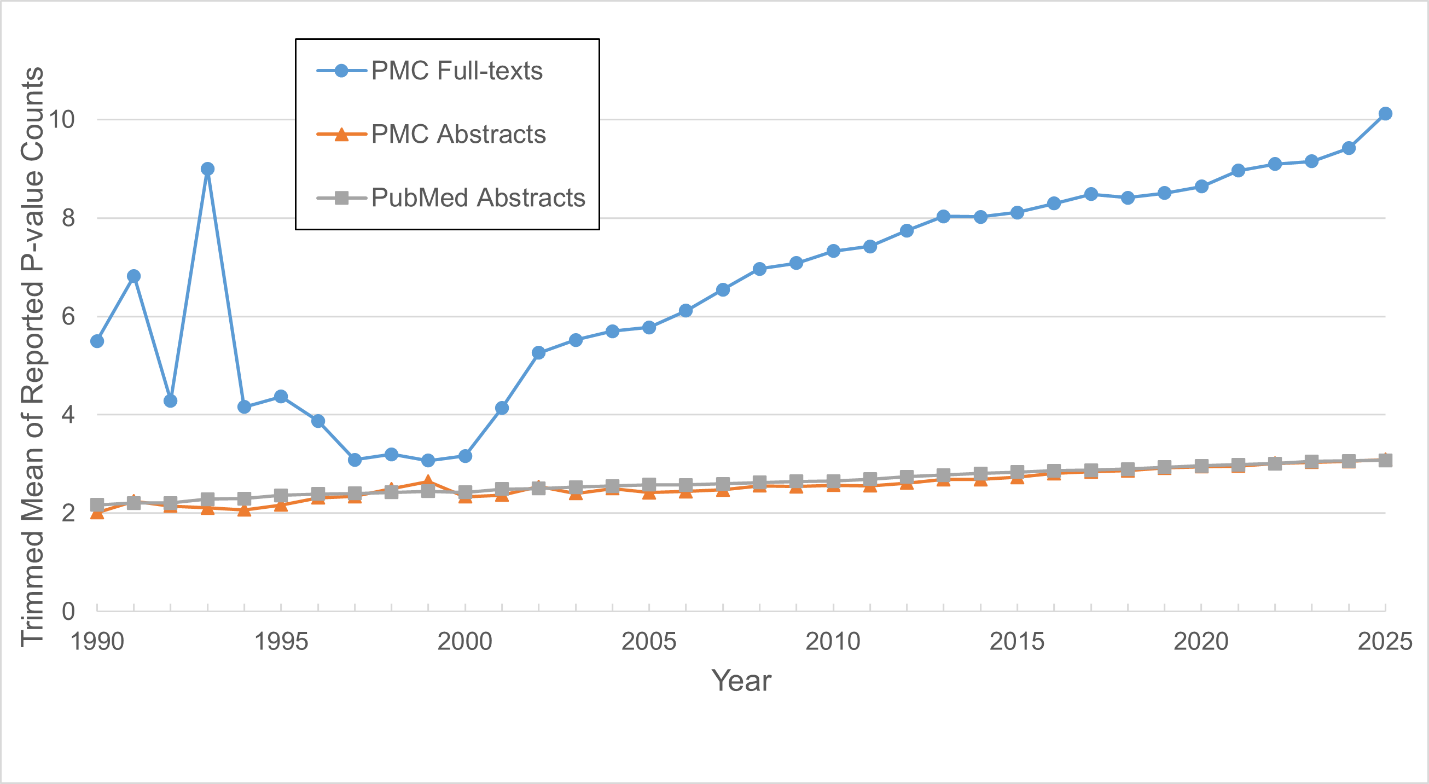

**Supplementary Figure 11. Trimmed Mean of Reported P-value counts per article by year.** The 5% trimmed mean (top/bottom 5% excluded) showed a similar upward trajectory. The proportions derived from PMC Full-texts data for the 1990–1996 interval may lack statistical stability, as this is the only period where the number of articles reporting at least one P-value is below 50.

**Data sharing**

The full code used in the study, from downloading items to printing results, is uploaded on GitHub Repository.

<https://github.com/mizoba/20250923PubMedStudy.git>

If you need additional information or would like to request the data used in the study, please contact us at the email below:

The full database is approximately 260 GB.
